# Immunogenicity and tolerability of booster typhoid conjugate vaccine (TCV) five to six years after initial dose in Burkinabe Children

**DOI:** 10.64898/2026.04.19.26351224

**Authors:** Jean W. Sawadogo, Alimatou Hema, Diarra Amidou, Jean Moise Kaboré, Denise Hien, Ludovic Kouraogo, Aminata R. Zou, Amidou Z. Ouédraogo, Alfred B. Tiono, Shrimati Datta, Marcela F. Pasetti, Kathleen M. Neuzil, Sodiomon B. Sirima, Alphonse Ouédraogo, Matthew B. Laurens

**Affiliations:** Groupe de Recherche Action en Santé (GRAS), Ouagadougou, Burkina Faso; Center for Vaccine Development and Global Health (CVD), University of Maryland School of Medicine, Baltimore, Maryland, USA; Gates Foundation, Seattle, Washington, USA

**Keywords:** Typhoid conjugate vaccine, booster dose, immunogenicity, seroconversion, Burkina Faso, vaccine safety, heterologous, public health

## Abstract

Typhoid fever remains a significant public health challenge in low- and middle-income countries. In 2018, The World Health Organization recommended a single dose typhoid conjugate vaccine (TCV) for routine immunization in endemic settings; however, evidence guiding booster doses remains limited. Homologous TCV booster doses have demonstrated immune boosting. This study assessed the immunogenicity and safety of a heterologous booster using a Vi capsular polysaccharide-CRM197 TCV (Vi-CRM) administered 5–6 years after primary vaccination with a Vi capsular polysaccharide tetanus toxoid TCV (Vi-TT) in children.

Children previously enrolled in a Phase 2 trial were recruited. Participants who had received TCV at 9–11 or 15–23 months were given a Vi-CRM booster at 6–7 years of age (Booster-TCV group), and controls received their first TCV dose at the same age (1st-TCV group). Serum anti-Vi IgG concentrations were measured at baseline and 28 days post-vaccination. Solicited and unsolicited adverse events (AEs) and serious adverse events (SAEs) were recorded.

Among 147 children enrolled, 87 received a second and 60 received a first TCV dose. Baseline anti-Vi IgG geometric mean titers (GMT) were higher in the Booster-TCV group (21.5 EU/mL; 95% CI: 17.2–26.8) than in the 1st-TCV group (5.5 EU/mL; 95% CI: 4.5–6.7). At day 28, GMTs rose markedly in both groups: 5140.0 EU/mL (95% CI: 4302.0–6141.3) in the Booster-TCV group and 2084.8 EU/mL (95% CI: 1724.4–2520.5) in the 1st-TCV group. Local reactions and systemic AEs were mild. No SAEs were observed. Vi-TT-induced immunity persisted for at least 5–6 years, and a heterologous booster triggered a strong immune response with universal seroconversion. These findings support heterologous prime–boost strategies to maintain protection in school-age children and inform optimization of TCV schedules in endemic regions.

## Introduction

Typhoid fever remains a major global public health challenge, particularly in low- and middle-income countries where access to safe water, sanitation, and healthcare is limited. Despite a marked decline in disease burden over the past three decades, estimates indicate that typhoid fever causes more than 7 million cases and approximately 93,000 deaths annually worldwide (1–3). Children, especially those under five years of age and school-aged, are disproportionately affected (4), underscoring the urgent need for effective preventive measures, including vaccination alongside improvements in water and sanitation.

Typhoid fever, caused by *Salmonella enterica* serovar Typhi, can lead to serious complications including intestinal perforation, gastrointestinal hemorrhage, and neurological complications in untreated cases with illness that lasts for more than two weeks (5). The ever-growing problem of antimicrobial resistance further complicates treatment (6) and can result in increased costs due to inadequate/unsuccessful treatment, a higher risk of complications, longer hospital stays, and likely higher mortality.

Typhoid conjugate vaccine (TCV) provides protection against *Salmonella* Typhi and is recommended by the World Health Organization (WHO) as a single dose vaccine for routine immunization in endemic areas. Compared with older polysaccharide vaccines (7), TCV offers important advantages including longer-lasting immunity and safe administration to infants as young as six months of age (8–10). Among TCV products, the Vi-tetanus toxoid conjugate vaccine (Vi-TT) and diphtheria toxin variant conjugate vaccine TYPHIBEV® Vi-CRM (Biological E., Ltd., Hyderabad, India), have demonstrated strong immunogenicity and safety in Africa and Asia (11–13). Nevertheless, data remain limited regarding optimal dosing schedules and the duration of protective immunity.

Multiple clinical trials have demonstrated TCV safety, immunogenicity, and efficacy, supporting vaccine introductions in multiple countries. Ultimately, these data, together with support from Gavi, the Vaccine Alliance, led to TCV being added to routine Expanded Programme on Immunisation (EPI) vaccination schedules for pediatric populations in endemic countries (8,9). Understanding both the duration of protection conferred by TCV and the need for and timing of booster doses is central to typhoid prevention and control. In Burkina Faso, an RCT demonstrated safety and immunogenicity of the Vi-TT TCV product administered with Expanded Program on Immunization (EPI) vaccines in children aged 9-11 and 15-23 months of age (8,9). This study cohort provided a unique opportunity to assess the safety and immunogenicity of a booster TCV dose with a different TCV product (Vi-CRM), administered approximately 5-6 years later. These data are essential for informing public health strategies to prevent typhoid fever and will add to existing data used by the WHO to inform TCV use within the Expanded Program on Immunization (EPI).

## Materials and Methods

### Study design

This clinical trial was conducted at a school-aged child outpatient clinic at Schiphra Protestant Hospital, an urban hospital in Ouagadougou, Burkina Faso. It follows a Phase 2 randomized, double-blind, controlled trial evaluating the safety and immunogenicity of Vi-TT among children younger than 2 years of age in Ouagadougou, and a subsequent assessment of the durability of vaccine-induced anti-Vi IgG and IgA responses in children immunized with TCV at the routine 15-month visit in Burkina Faso (NCT03614533) (8–10).

### Participants

Details of the original studies are published (8,9). In brief, we enrolled and vaccinated two cohorts of healthy children: 100 aged 9–11 months and 150 aged 15–23 months. The younger cohort was randomized 1:1 to receive either Vi-TT (Group 1) or an inactivated polio vaccine (IPV; Group 2) as the control. In the older cohort, participants were randomized into one of three groups: Vi-TT and IPV with delayed meningococcal A conjugate vaccine (MCV-A, Group 1), Vi-TT and MCV-A (Group 2), or MCV-A and IPV (Group 3). All vaccines were administered intramuscularly along with routine measles-rubella (MR) and yellow fever (YF) vaccines.

This study re-enrolled children who had previously participated in the phase 2 trial in Ouagadougou. In the current study, participants who originally received Vi-TT were administered a second dose of TCV using the Vi-CRM product, while those who had not previously received TCV were given their first dose at 6–7 years of age using the Vi-CRM product. The primary objective was to assess and compare the immune response elicited by a first dose of TCV administered at 6–7 years with that elicited by a booster dose in children previously vaccinated with TCV.

Families of children who had previously participated in the TCV study were contacted by phone call or via community-based health workers serving as guides. Detailed study information was provided to these families through individual or group discussions. Parents or guardians received both oral and written explanations, in local and French languages, of study procedures and provided written informed consent in a private setting before enrollment and blood sample collection.

Inclusion criteria required that participants and their parent or legal guardian reside within the study area and provide informed consent. Eligible participants included male and female children enrolled in the 2018-2019 Phase 2 trial. Participants were excluded if they received blood products within the previous six months, received TCV outside the study (e.g., through a vaccination campaign or routine immunization), or had any condition deemed by the investigator to potentially interfere with vaccine evaluation or pose a health risk. Additionally, temporary exclusion criteria were applied to participants who met any of the following conditions, with reassessment after 48 hours: reported fever within 24 hours before vaccination and/or use of any antipyretic medications within 4 hours before vaccination.

### Randomization and masking

This study did not involve randomization, as participants were assigned based on predefined criteria according to study objectives. The TCV vaccination status of participants from the initial study remained blinded to study personnel except for the study biostatistician throughout this trial.

### Procedures

TYPHIBEV® Vi-CRM (Biological E., Ltd., Hyderabad, India), contains 25 μg of Vi polysaccharide conjugated to a protein carrier, which is a variant of diphtheria toxin (CRM197). Vi-CRM was administered intramuscularly in the left upper deltoid. Vaccinations were performed at the outpatient clinic of Schiphra Protestant Hospital in Ouagadougou, Burkina Faso. Trained local study personnel not involved in participant follow-up or adverse event (AE) monitoring administered the injections. On study Day 0, each participant underwent blood sampling and subsequently received the Vi-CRM vaccine. After vaccination, participants were monitored onsite for serious reactions, and via phone on Day 7 post-vaccination to inquire about local and systemic AEs that occurred each day since vaccination. Study personnel were available for unscheduled visits during the 28-day follow-up period. Unsolicited AEs and serious AEs (SAEs) were recorded until Day 28. Adverse events were graded according to the United States Food and Drug Administration (FDA) guidelines for vaccine clinical trials (US Food and Drug Administration, 2007). Blood samples were collected for immunogenicity assessment on Days 0 and 28. Anti-Vi IgG antibody responses were measured using the VaccZyme Salmonella Typhi Vi Immunoglobulin G (IgG) ELISA kit (The Binding Site Group Ltd, Birmingham, UK). Data analysis was performed at the University of Maryland, Baltimore in Baltimore, Maryland, United States of America.

### Outcomes

The primary outcome was the immunogenicity of a first TCV dose administered at 6–7 years of age compared with a second dose in children who previously received TCV at 9–11 or 15–23 months of age. Immunogenicity was assessed by measuring anti-Vi IgG antibody responses before and 28 days after vaccination. Seroconversion was defined as a 4-fold rise in anti-Vi IgG titers from baseline to Day 28.

Secondary outcomes included TCV safety and reactogenicity, assessed via solicited and unsolicited AEs. Safety assessments were conducted through active and passive surveillance. Local and systemic adverse events (AEs) were recorded for 7 days after vaccination. Additionally, unsolicited AEs were documented, and any SAEs were carefully monitored throughout the study period to ensure participant safety.

### Statistical analysis

All data were analyzed with SAS software Version 9.4 (Copyright ® 2013 SAS Institute Inc., Cary, NC, USA).

### Sample size determination

A total of 250 participants were initially divided into two groups: 148 received Vi-TT, and 102 received IPV. Accounting for a 25% failure rate in contacting and recruiting participants, 187 children (75% of 250 enrollees) were expected to complete the follow-up assessments.

### Analysis sets

Descriptive statistics were used to summarize baseline demographics, with age presented as mean ± standard deviation and sex as proportions. Safety analyses followed an intention-to-treat (ITT) approach, including all participants with available post-vaccination safety data. The incidence of local and systemic AEs within seven days post-vaccination was compared between the Booster-TCV and 1st-TCV groups. The incidence of unsolicited AEs and SAEs within 28 days of vaccination were also compared between the same groups.

Immunogenicity was evaluated in the ITT population, comprising participants with samples collected at day 28 and 30–35 months post-vaccination. Anti-Vi IgG titers were log-transformed and compared within each group using paired t-tests and between each group using two sample t-test with unequal variances. Antibody distributions were visualized and reported using sample size, geometric mean titers, 95% confidence intervals, and p-values.

A multi-time point immunogenicity analysis was conducted within the ITT population, restricted to participants enrolled in the sub-study assessing long-term anti-Vi IgG responses. A previous investigation assessed anti-Vi IgG and IgA antibodies 30–35 months after vaccination in a subset of participants from a clinical trial in which TCV had been co-administered with the routine vaccine at the 15-month vaccination visit (10). In the present analysis, antibody titers were compared from baseline to 30–35 months after primary TCV administration, and from baseline to 28 days post-booster dosing at 6–7 years.

## Results

### Baseline Characteristics

Of 250 children originally vaccinated between 9 and 23 months of age in the randomized controlled trials, 157 (62.8%) were successfully recontacted in 2024, and 152 were deemed eligible for enrolment. Following five refusals, 147 participants were vaccinated between August and October 2024: 87 in the Booster-TCV arm and 60 in the 1st-TCV arm. The two arms showed comparable demographic profiles, with a mean age of 6.8 years (SD ±0.3) and similar gender distributions—57.5% male in the Booster-TCV arm and 45.0% in the 1st-TCV arm. Among the 147, 88 children (59.9%) were in the durable anti-Vi IgG sub-study, with 52 in the Booster-TCV group and 36 in the 1st-TCV group (Figure 1 and Table 1).

**Figure 1.**
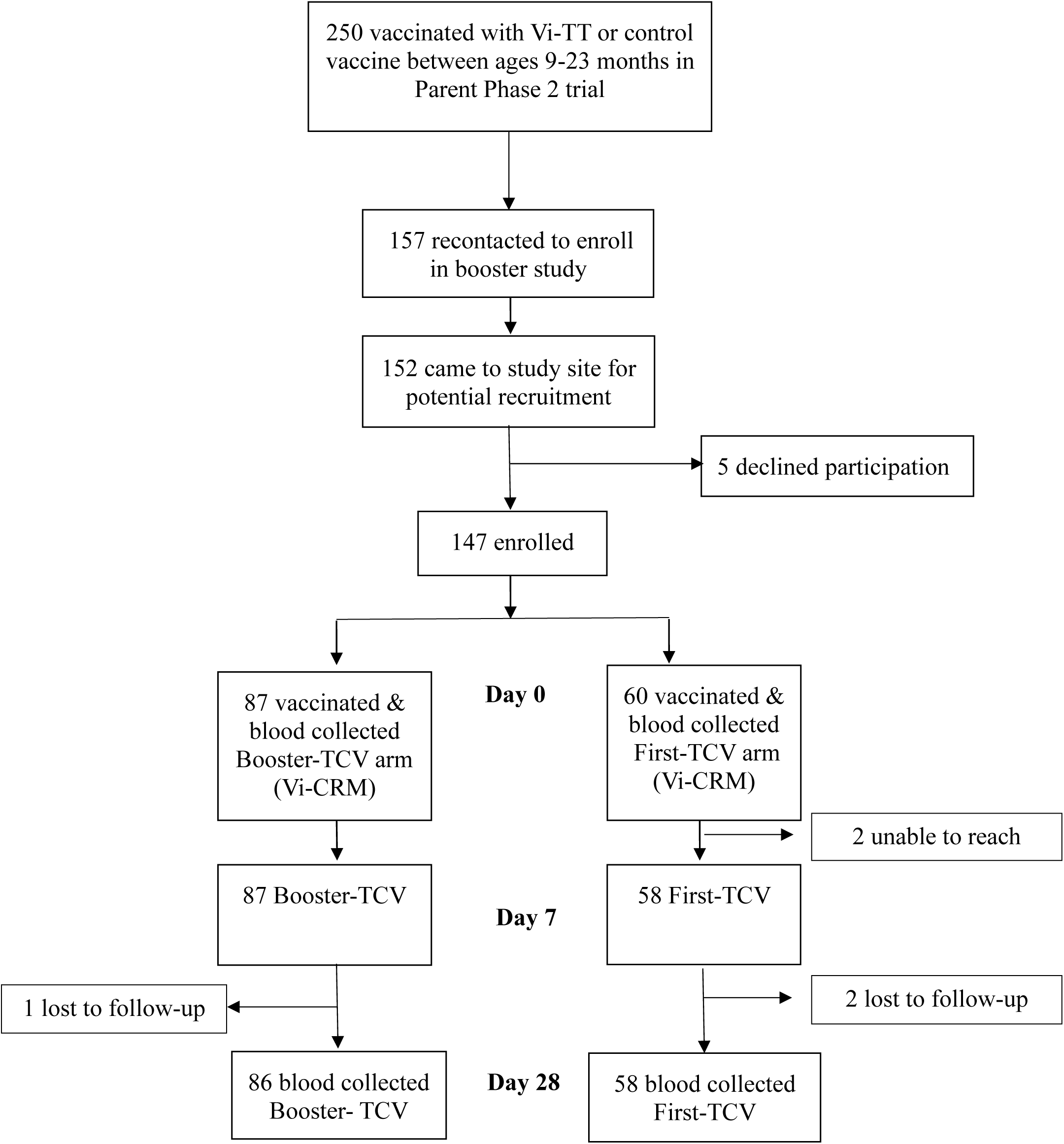
Flow of participant enrollment, allocation, and follow-up in the typhoid conjugate vaccine booster study.

**Table 1:** Baseline socio-demographic characteristics.

|  | **Booster TCV**<br>**n=87** | **First TCV**<br>**n=60** | **Overall**<br>**n=147** |
| --- | --- | --- | --- |
| **Age at enrolment**<br>**Mean (±SD)** | 6.8 (0.3) | 6.7 (0.3) | 6.8 (0.3) |
| **Sex** |  |  |  |
| **Female n (%)** | 37 (42.5) | 33 (55.0) | 70 (47.6) |
| **Male n (%)** | 50 (57.5) | 27 (45.0) | 77 (52.4) |

### Safety profile

Local injection site reactions within seven days post-vaccination were reported exclusively in the Booster-TCV group, affecting 4.6% of participants (95% CI: 1.3–11.4, Table 2). These reactions were mild pain or tenderness at the injection site. No local reactions were observed in the 1st-TCV group.

**Table 2.** Summary of reactogenicity and safety parameters (adverse events) by vaccine arm in the intention to treat population after Vi-CRM vaccination.

|  |  | **Booster-TCV**<br>**n=87** | **1st-TCV**<br>**n=58** |
| --- | --- | --- | --- |
| **Local Reactions at injection site (Day 0-7)** |  |  |  |
|  | **Pain/tenderness** | 4 (4.6, 1.3-11.4)<br>Mild (4) on Day 1 | 0 (0.0, 0.0-6.2) |
|  | **Swelling** | 0 (0.0, 0.0-4.2) | 0 (0.0, 0.0-6.2) |
|  | **Erythema** | 0 (0.0, 0.0-4.2) | 0 (0.0, 0.0-6.2) |
|  | **Any local reaction** | 4 (4.6 1.3-11.4) | 0 (0.0, 0.0-6.2) |
| **Systemic Reactions (Day 0-7)** |  |  |  |
|  | **Fever** | 2 (2.3, 0.3-8.1)<br>Day 2 (1), Day 6 (1) | 4 (6.9, 1.9-16.7)<br>Day 2 (1), Day 3 (3) |
|  | **Feverishness** | 0 (0.0, 0.0-4.2) | 0 (0.0, 0.0-6.2) |
|  | **Myalgia** | 0 (0.0, 0.0-4.2) | 0 (0.0, 0.0-6.2) |
|  | **Arthralgia** | 0 (0.0, 0.0-4.2) | 0 (0.0, 0.0-6.2) |
|  | **Malaise** | 0 (0.0, 0.0-4.2) | 0 (0.0, 0.0-6.2) |
|  | **Any systemic reaction** | 2 (2.3, 0.3-8.1) | 4 (6.9, 1.9-16.7) |
| **Unsolicited adverse events (Day 0-28)** |  |  |  |
|  | **Related** | 0 (0.0, 0.0-4.2) | 1 (1.7, 0.04-8.9) |
|  | **Not related** | 23 (26.4, 17.6-37.0) | 18 (30.0, 18.9-43.2) |
|  | **Any unsolicited adverse event** | 23 (26.4, 17.6-37.0) | 19 (31.7, 20.3-45.0) |
| Data are n (% , 95% CI). n=number of participants. CI=confidence interval. Vi-CRM: Vi polysaccharide diphtheria toxin conjugated vaccine. |  |  |  |

Systemic AEs were slightly more frequent in the 1st-TCV group, with an incidence of 6.9% (95% CI: 1.9–16.7), compared to 2.3% (95% CI: 0.3–8.1) in the Booster-TCV group (Table 2), but this difference was not statistically significant. Reported fever was the only systemic AE consistently reported, occurring in four participants from the 1st-TCV arm. No other systemic symptoms were noted in either group.

All reported adverse events resolved without sequelae by day 28, and no serious adverse events related to the vaccine were observed.

Unsolicited adverse events (AEs) occurring after vaccination were reported at comparable rates in both study arms: 26.4% (95% CI: 17.6–37.0) in the Booster-TCV group and 31.7% (95% CI: 20.3–45.0) in the 1st-TCV group. Most events were mild or moderate, and none were vaccine-related, with rhinitis and uncomplicated malaria being the most frequently reported conditions (Supplementary Table 1).

No serious adverse event (SAE) was recorded during the study. All unsolicited AEs resolved without sequelae.

### Immunogenicity

#### IgG Geometric Mean Titer and Seroconversion at Day 28 After Vaccination

At baseline just before second vaccinations, children in the Booster-TCV arm exhibited significantly higher anti-Vi IgG geometric mean titers (GMT) compared to those in the 1st-TCV arm (21.5 EU/mL; 95% CI: 17.2–26.8 vs. 5.5 EU/mL; 95% CI: 4.5–6.7), reflecting sustained antibody levels from prior immunization. By day 28 post-vaccination, both groups demonstrated marked increases in GMTs, with the Booster-TCV arm reaching 5140.0 EU/mL (95% CI: 4302.0–6141.3), significantly higher than the 1st-TCV arm, which reached 2084.8 EU/mL (95% CI: 1724.4–2520.5) (Figure 2).

**Figure 2:**
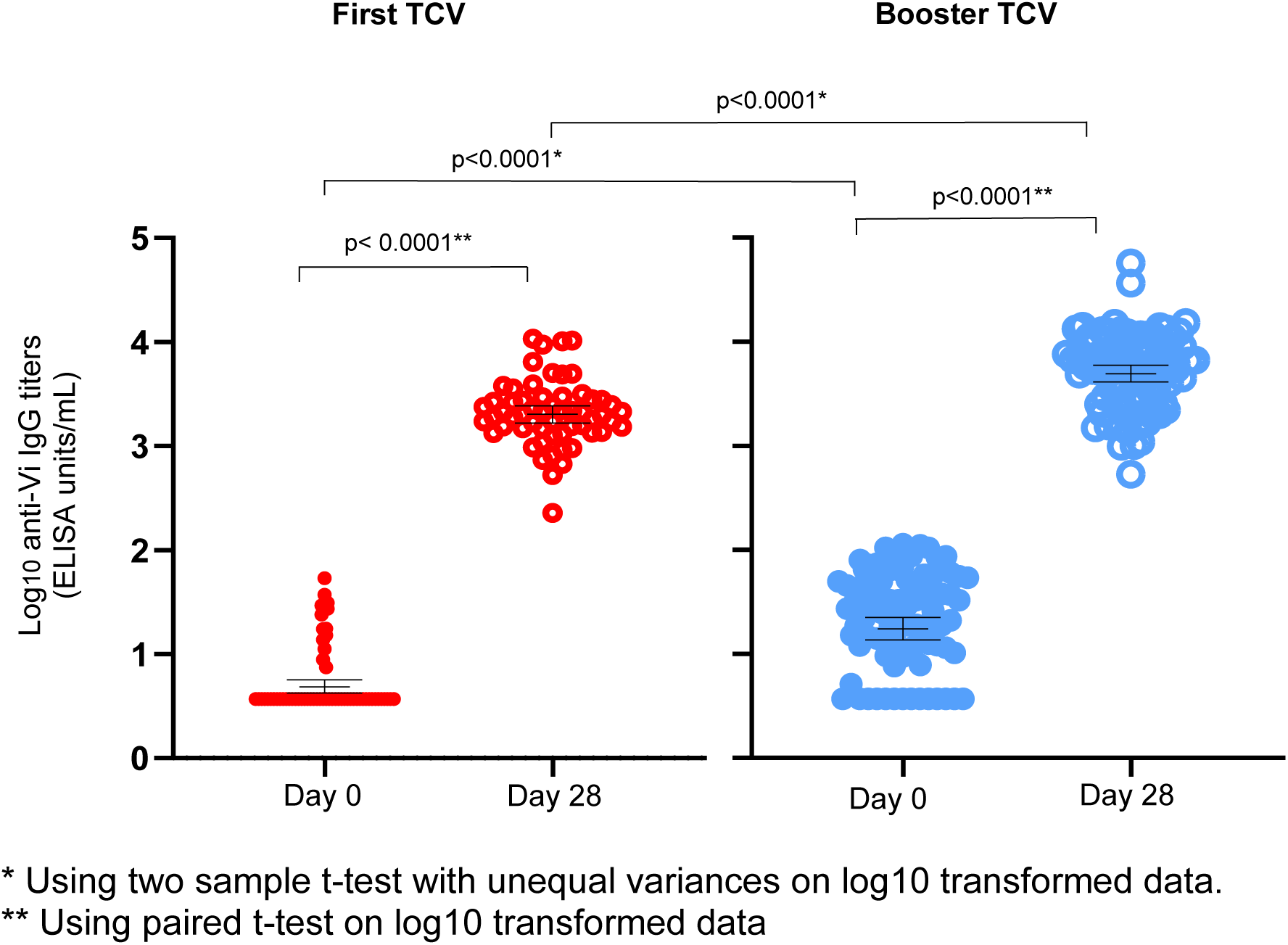
Anti-Vi IgG before and 28 days after Vi-CRM vaccination in the intention-to-treat population.

Seroconversion, defined as a ≥4-fold rise in anti-Vi IgG titers, was observed in all participants across both arms by day 28, with one exception in the 1^st^ TCV group (Table 4).

**Table 4:**
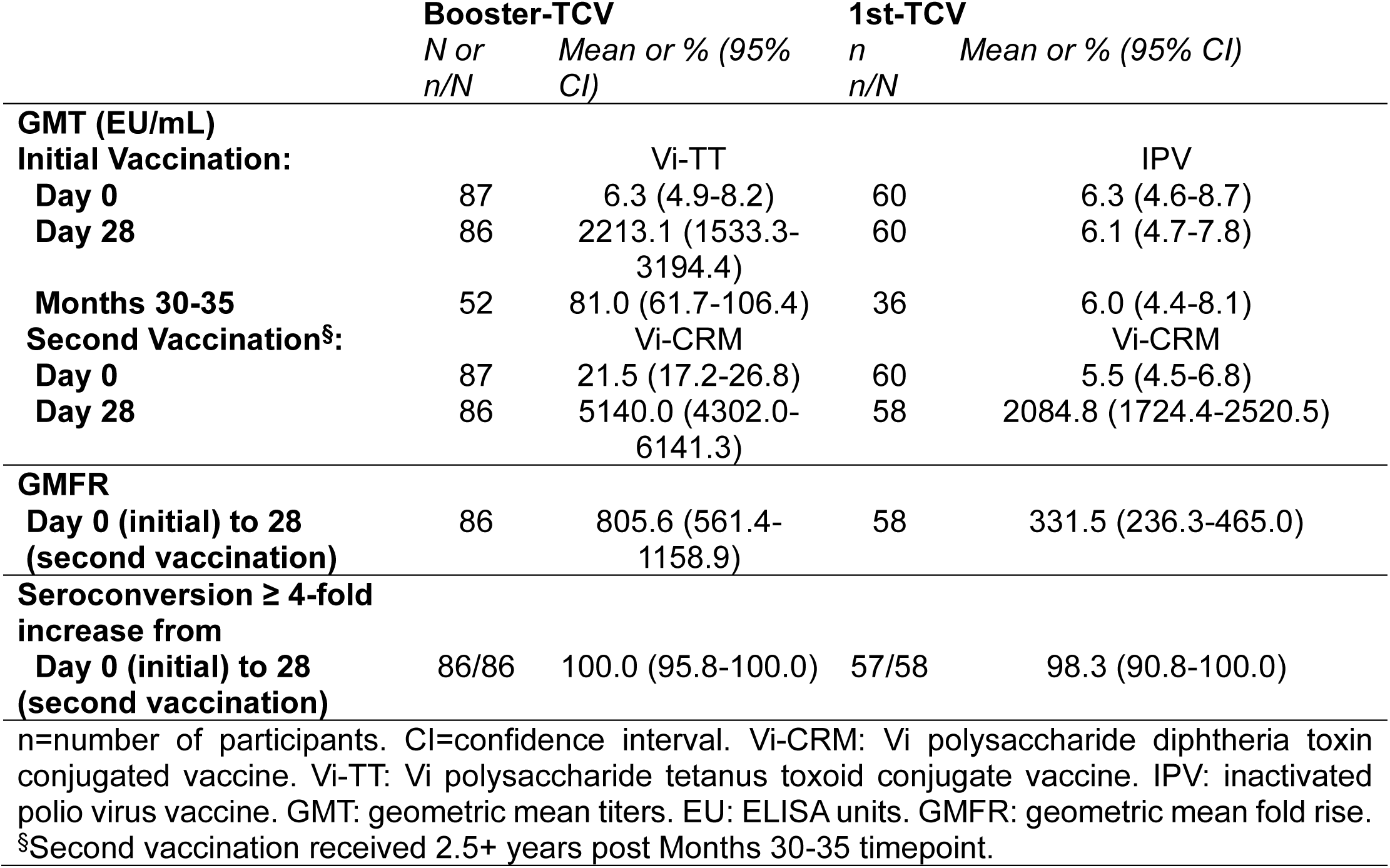
Anti-Vi IgG before and after first IPV or Vi-TT vaccination and before and after Vi-CRM vaccination in the intention to treat population.

|  | **Booster-TCV** |  | **1st-TCV** |  |
| --- | --- | --- | --- | --- |
|  | *N or*<br>*n/N* | *Mean or % (95%*<br>*CI)* | *n*<br>*n/N* | *Mean or % (95% CI)* |
| **GMT (EU/mL)** |  |  |  |  |
| **Initial Vaccination:** |  |  |  |  |
|  |  | Vi-TT |  | IPV |
| **Day 0** | 87 | 6.3 (4.9-8.2) | 60 | 6.3 (4.6-8.7) |
| **Day 28** | 86 | 2213.1 (1533.3-3194.4) | 60 | 6.1 (4.7-7.8) |
| **Months 30-35** | 52 | 81.0 (61.7-106.4) | 36 | 6.0 (4.4-8.1) |
| **Second Vaccination<sup>§</sup>:** |  |  |  |  |
|  |  | Vi-CRM |  | Vi-CRM |
| **Day 0** | 87 | 21.5 (17.2-26.8) | 60 | 5.5 (4.5-6.8) |
| **Day 28** | 86 | 5140.0 (4302.0-6141.3) | 58 | 2084.8 (1724.4-2520.5) |
| **GMFR** |  |  |  |  |
| **Day 0 (initial) to 28 (second vaccination)** | 86 | 805.6 (561.4-1158.9) | 58 | 331.5 (236.3-465.0) |
| **Seroconversion ≥ 4-fold increase from** |  |  |  |  |
| **Day 0 (initial) to 28 (second vaccination)** | 86/86 | 100.0 (95.8-100.0) | 57/58 | 98.3 (90.8-100.0) |
| n=number of participants. CI=confidence interval. Vi-CRM: Vi polysaccharide diphtheria toxin conjugated vaccine. Vi-TT: Vi polysaccharide tetanus toxoid conjugate vaccine. IPV: inactivated polio virus vaccine. GMT: geometric mean titers. EU: ELISA units. GMFR: geometric mean fold rise. |  |  |  |  |
| <sup>§</sup>Second vaccination received 2.5+ years post Months 30-35 timepoint. |  |  |  |  |

#### IgG Geometric Mean Titer and Seroconversion at multi-time point over vaccination

As shown in Figure 3 and Table 4, a multi-time point analysis of anti-Vi IgG responses was conducted among participants in the durable antibody sub-study of the randomized controlled trial. This analysis captured immune responses after initial Vi-TT vaccination at 9 or 15 months of age through post-booster administration at 6–7 years. At baseline (before the initial Vi-TT vaccination), anti-Vi IgG GMTs were comparable between the Booster-TCV and 1st-TCV groups. After primary vaccination, GMTs in the Booster-TCV group rose significantly to 2213.1 EU/mL (95% CI: 1533.3–3194.4) by day 28. A gradual decline was observed over time, with GMTs decreasing to 81.0 EU/mL (95% CI: 61.7–106.4) at the 30–35-month timepoint. At enrolment into the booster study, GMTs in the Booster-TCV group were 21.5 EU/mL (95% CI: 17.2–26.8), indicating persistence of antibody responses. Post-booster, anti-Vi IgG GMTs increased markedly to 5140.0 EU/mL (95% CI: 4302.0–6141.3) by day 28, demonstrating a strong recall response. In the 1st-TCV group, anti-Vi IgG GMTs remained low from infancy until the first TCV dose at age 6-7 years, after which titers rose to 2084.8 EU/mL (95% CI: 1724.4–2520.5) by day 28.

**Figure 3:**
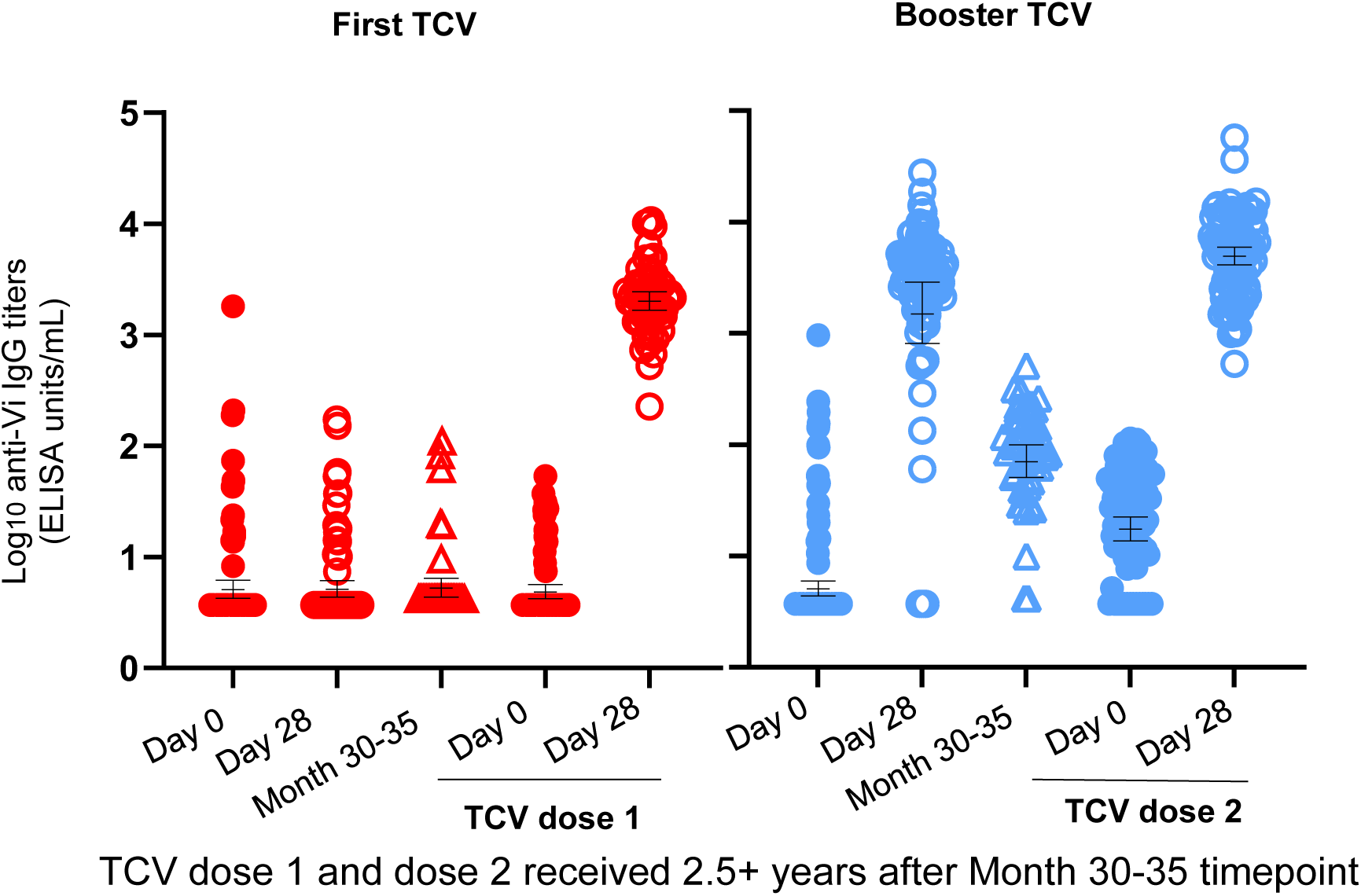
Anti-Vi IgG before and after the first IPV or Vi-TT vaccination and before and after Vi-CRM vaccination in the intention-to-treat population.

## Discussion

In this study, we assessed the persistence of anti-Vi IgG responses 5–6 years after primary TCV administration in early childhood, and evaluated the immunogenicity and safety of a heterologous booster dose delivered at school age. Our findings show that vaccine-induced immunity remains detectable years after initial vaccination, and that a booster dose elicits a strong recall response, with universal seroconversion and a favorable safety profile. These results provide important evidence on the durability of TCV-induced protection in children living in a typhoid-endemic setting and offer timely insights to inform policy discussions on the potential role of booster dosing to sustain immunity where typhoid burden remains high.

### Immunogenicity Findings in Context of Prior Evidence

The immunogenicity results should be interpreted in the broader context of emerging evidence on the durability of long-term TCV responses. While TCVs elicit robust initial protection, emerging data suggest that long-term efficacy may vary by age at first vaccination. A RCT in Malawi reported approximately 80% efficacy of a single TCV dose sustained over at least four years (14). However, data indicated a nonsignificant trend toward reduced efficacy and more pronounced antibody waning in children vaccinated before two years of age (14,15). A randomized controlled trial (RCT) in Malawi demonstrated that a single dose of TCV provided over 78% efficacy against blood-culture confirmed typhoid fever for at least four years (16). However, children vaccinated before the age of two exhibited greater waning in antibody responses over time compared to older children. Although these differences were not statistically significant, they raise the possibility that booster dosing may be particularly relevant for children vaccinated before age two.

Findings from other settings further highlight this heterogeneity. Long-term data from the Philippines indicate that a TCV booster dose sustained protective immune responses for at least five years, supporting its potential role in durable typhoid prevention (17). In contrast, data from Bangladesh indicate a more rapid decline in vaccine-induced protection within 3 to 5 years after single dose TCV administration (18). Together, these observations underscore the need to further study the longevity of TCV efficacy across different age groups and epidemiologic contexts, and the potential role of booster dosing in endemic areas.

Additional evidence from Malawi demonstrated that a second dose of Vi-TT administered at 5 years of age following primary vaccination at 9-11 months was well tolerated and elicited a significantly greater immune response than a first dose at 5 years (16). The magnitude of the booster response observed in that study closely parallels that observed in the present study conducted in Burkina Faso using a heterologous Vi-CRM booster. At 28 days post-boost, immune responses were comparable, including anti-Vi IgG titer (6867.9 EU/mL, 95% CI 5794.1-8140.6), geometric mean fold rise (375.7, 95% CI 293.2-484.4) and seroconversion (100%, 95% CI 94.8-100.0)(16), supporting the interchangeability of homologous and heterologous booster approaches. These findings provide empirical support for the interchangeability of licensed TCV products for booster dosing.

The durability of antibody responses observed 5-6 years after primary vaccination is consistent with established immunological principles. Immune responses in infants are often less durable than those generated in later childhood, reflecting developmental differences in B-cell maturation, germinal center reactions, and memory formation (19–21). Previous work has demonstrated that immune responses to polysaccharide antigens, including Vi, vary substantially by age, and that conjugate vaccines only partially overcome these developmental constraints. Our demonstration of sustained anti-Vi IgG responses several years after primary vaccination in children immunized between 9 and 23 months of age adds important longitudinal data to the growing body of evidence on TCV durability.

The marked increase in antibody titers after revaccination aligns well with established principles of immunological memory induced by conjugate vaccines. Protein-polysaccharide conjugate vaccines prime memory B cells that can rapidly expand and differentiate upon antigen re-exposure, generating higher antibody concentrations than those achieved by primary vaccination (22). This phenomenon has been consistently observed with meningococcal, pneumococcal, and typhoid conjugate vaccines (16,23,24), and is reinforced by the strong recall responses documented in the present study.

### Safety Profile in Broader Context

The safety profile observed in this study is consistent with extensive evidence supporting the favorable tolerability of TCVs across pediatric age groups (8,9,13). Both solicited and unsolicited adverse events were predominantly mild and transient, and no vaccine-related SAEs were identified. These findings support the safety of TCV administration as both a primary and booster dose in school-aged children and expand the global safety database for TCVs, which is informed by clinical trials and post-introduction surveillance from multiple countries (25–27).

The reactogenicity profile observed in this study, characterized by mostly mild local reactions and minimal systemic symptoms, is typical of protein-conjugate vaccines (16,28,29). Previous studies have similarly reported acceptable safety profiles for TCVs administered as either primary or booster doses across age groups (30,31). Importantly, the present study specifically addresses a gap in the literature by demonstrating the safety of long-interval booster dosing administered 5 to 6 years after primary vaccination.

### Implications for Vaccination Policy

These findings have important implications for typhoid vaccination strategies in endemic settings. WHO’s Strategic Advisory Group of Experts (SAGE) on Immunization recently reviewed the longer-term immunogenicity and efficacy data and endorsed that countries consider introducing a booster dose around 5 years of age in settings with very high typhoid incidence for children who received a primary TCV dose at 9–24 months of age (32). In other incidence settings where the primary dose of TCV is administered before 24 months of age, a booster dose may be considered if evidence of waning protection is observed in vaccinated cohorts, especially in areas with a high case fatality ratio or high prevalence of antimicrobial resistance (32). Final WHO recommendations will be released in an upcoming Weekly Epidemiological Record. Modeling studies suggest that maximizing coverage with a single dose may yield greater immediate public health impact than routine booster deployment, particularly in high-incidence settings. At the same time, economic and epidemiologic assessments emphasize the need for context-specific cost-effectiveness analyses when considering booster strategies (33). If boosters are adopted, delivery through established platforms such as school-based programs could minimize logistical challenges (34,35), and the demonstrated interchangeability of TCV products offers additional programmatic flexibility.

### Heterologous TCV vaccination strategies and Public Health Significance

In this study, we evaluated a heterologous TCV booster strategy in children primed with Vi–TT before age two and boosted with Vi–CRM at school age, and compared immune responses to those elicited by a single dose of Vi-CRM administered at school age. Baseline anti-Vi IgG levels were higher in booster recipients, indicating sustained immune memory. Post-vaccination, both groups exhibited strong antibody responses, with higher titers observed in the booster group and universal seroconversion in both groups. Safety outcomes were comparable, with no related SAEs.

These findings support the feasibility and immunological benefit of a heterologous prime–boost approach to enhance immunogenicity and extend protection at school entry in typhoid-endemic regions. Given the global burden of typhoid, especially among children in South Asia and sub-Saharan Africa, optimizing the timing and delivery of TCV schedules remains a critical public health priority (36).

## Acknowledgements

We extend our sincere gratitude to the study participants and their families for their invaluable contribution. We also acknowledge the dedicated teams at Groupe de Recherche Action en Santé (GRAS) and Schiphra Protestant Hospital (Ouagadougou), the Center for Vaccine Development and Global Health, and the Clinical and Translational Research Informatics Center at the University of Maryland School of Medicine.

We thank Biological E. Limited for providing the investigational vaccine used in this study. This study was funded in part by a grant from the Gates Foundation (INV-030857). The funders had no role in study design, data collection and analysis, decision to publish, or preparation of the manuscript.

## Data Availability Statement

Individual participant data underlying the results reported in this article, after de-identification, will be made available. Data will be accessible through the Vivli Center for Global Clinical Research Data platform (https://vivli.org/) to researchers who provide a methodologically sound proposal, following approval of a data access request and execution of a data use agreement.

